# Memory for Music (M4M) protocol for an international randomized controlled trial: Effects of individual intensive musical training based on singing in non-musicians with Alzheimer’s disease

**DOI:** 10.1101/2024.09.25.24313991

**Authors:** Marcela Lichtensztejn, Anja-Xiaoxing Cui, Monika Geretsegger, Astri J. Lundervold, Stefan Koelsch, Daniela M. Pfabigan, Jörg Assmus, Elias Langeland, Carolina Tabernig, Ragnhild E. Skogseth, Christian Gold

## Abstract

**Introduction:** The number of people living with dementia is increasing worldwide. Alzheimer’s disease (AD) is the most common type of dementia. It typically manifests itself initially with cognitive impairment in the memory domain, and gradually progresses towards affecting all activities of daily living. Active music interventions, particularly singing, may improve mood, social behaviour, and quality of life. However, little is known about their effects on cognition, although some studies have provided promising results. The M4M project aims to fill this gap in research by measuring the effects of learning new songs on cognitive functioning. Specifically, M4M will examine memory for new songs in non-musician adults with AD after undergoing intensive versus minimal individual musical training based on singing novel songs.

**Methods and analysis:** Home-dwelling adults with AD, 65 years or older, will receive 5 months of intensive intervention (2x/week) and 5 months of minimal intervention (1x/month). In a crossover design, participants will be randomized to receive either the intensive or minimal intervention first, with 2 months between the intervention periods. Participants will receive individual music lessons to learn new songs, provided by a music instructor with adequate training. The main outcomes will be measured at the beginning and end of each intervention period. General cognition will be measured with the Alzheimer’s Disease Assessment Scale – Cognitive (ADAS-cog) by an assessor blinded to the randomisation. Participants’ memory for music will be measured using the N400 component of electroencephalographic event-related potentials in response to music stimuli. Additional outcomes evaluated during intervention sessions include mood and musical performance observations. With 113 participants randomised, the trial will have 80% power to detect clinically meaningful effects. Relations between mood, memory for music, and cognitive abilities will be examined, with sex, age, AD stage, previous musical training, and education as covariates. M4M will be conducted in close collaboration between academic researchers, service providers, and service users to ensure relevance and applicability.

**Ethics and dissemination:** Dissemination of findings will apply to local, national, and international levels. The study has been approved by the Regional Committees for Medical and Health Research Ethics in Norway.

**Trial registration:** ClinicalTrials.gov, NCT06611878.

**Strength and limitations:**

- Based on recent data suggesting that individuals with advanced dementia can learn new songs, our study moves beyond reminiscence-based therapy since the participants will be actively involved in musical training based on singing to learn novel songs
- The study focuses on home-dwelling older adults with dementia. This is becoming more and more important, as interventions that can prolong the period of independent living outside care facilities are urgently needed
- By using EEG technology that is portable, inexpensive, non-invasive, less demanding for participants than other brain imaging examinations, performed in a naturalistic setting, this study will reach people who are less mobile or live in remote areas, thus improving generalisability.
- As a multinational trial conducted in urban and rural settings in high- and middle-income countries, results will be relevant across diverse societies.
- Due to the nature of the intervention, participants cannot be blinded.

## INTRODUCTION

There are 55 million people living with dementia worldwide, and this number is expected to increase to 78 million by 2030 and 139 million by 2050 (1). As the most common form of dementia, Alzheimer’s disease (AD) contributes to 60-80% of cases (2). AD is a neurodegenerative disease which commonly presents with cognitive impairment in the memory domain, with a gradual affection of other cognitive, social, and emotional domains. Early cognitive impairments often affect episodic memory, naming, semantic memory, and verbal fluency (3). As the disease progresses, the person tends to experience increasing disorientation in time and space, changes in sleep patterns and mood, significant personality changes, communication difficulties, and motor disorders. This cluster of symptoms eventually leads to loss of independence, with an increasing need for assistance in activities of daily living (2).

Although cognitive symptoms typically appear years after the pathology is established in the brain (4), cognitive complaints are crucial for clinical attention. AD is characterized by extracellular amyloid beta plaques and intracellular neurofibrillary tangles of hyperphosphorylated tau protein. These proteins disrupt the communication between neurons, leading to neuronal loss and altered rhythmic patterns (5). Memory problems in AD and other forms of dementia are closely linked to changes in the hippocampus, which relates to the impairment to encode new information and to retrieve it later. These biochemical changes in the hippocampus later spread to other temporal regions (5).

Identifying and providing suitable interventions is essential to improve the quality of life of those with AD and may delay the progression of the disease. Music interventions (6), particularly those involving singing (7), have shown a positive impact on mood, behaviour and quality of life of people with dementia. Additionally, active music-making is positively associated with several cognitive functions, including learning and memory (8).

People with AD, whether they are musicians or not, tend to have a well-preserved musical semantic memory for songs learned prior to the onset of the disease, which may contrast sharply with their general cognitive functioning deficits (9–12). Musical semantic memory refers to “known” melodies, that is, purely musical information stored and accessed independently (independent evocation evidenced by singing) or by sense of familiarity when listening to a melodic progression (recognition of the melody of a piece of music), regardless of the timbre or tonality, stripped of any non-musical contextual information (such as title, name of the composer, musical era to which it belongs, past event in which this melody was heard, etc.). Therefore, this conception for musical memory involves the retention of musical information without the associated non-musical details (13).

However, individuals with AD exhibit lower performance when becoming familiar with new music or when learning new songs compared to healthy non-musician adults. Healthy non-musician adults have been shown to recognize new musical material 24 hours later after being exposed three times to a list of 24 unknown musical extracts under three different conditions (with one minute pause between each time): after the intake of a dopaminergic antagonist, a dopaminergic precursor, and a placebo (14). Non-musician adults with AD in a moderate to severe stage are shown to need more than 3 repetitions for new items to display explicit encoding abilities. When repeatedly exposed to listen to instrumental music, they may need four sessions per week for 2 weeks to develop a sense of familiarity (15), or they may become familiar with a novel 10-line song in 8 weeks by being exposed to learn it during once a week singing workshops (16). However, there has been a case of a non-musician adult with moderate to advanced AD who displayed an outstanding and increasingly improved ability to learn new songs over four years by attending individual adapted music lessons based on singing. This improvement occurred despite an overall deterioration due to the progression of the disease (17).

Altered brain structure and function is well documented in patients with AD, and several markers have been suggested. Among others, event-related potentials (ERPs) assessed via EEG recordings can be used to study aspects of how the brain processes information in people with AD (5). It has been consistently reported that adults with AD show impairments in the N400 component during tasks assessing lexical and linguistic semantic processing (5). The N400 component is a negative ERP deflection peaking between 250–550ms after stimulus onset, with maximal distribution over the centro-parietal electrode sites. It is commonly observed when semantic expectations are violated, and presumably originates from posterior middle temporal and parahippocampal gyri (5,18). Its amplitude has been shown to be reduced and delayed in healthy elderly adults (19), with even smaller amplitude or prolonged latencies and altered topography in response to linguistic stimuli (20) in those with AD compared to healthy controls (5). This likely reflects dysfunction of semantic memory processes.

An N400-like component (we will refer to it as “N400” for simplicity) can also be elicited by non-verbal stimuli, for example by violations of expected notes in a melody. Several studies have used the N400 component to investigate the processing of meaning with music. In addition, one study has investigated musical memory using the N400, suggesting that the N400 may also serve as a marker of musical memory, although its timing and origin is less solidly established than in response to verbal language stimuli (21–23). Previous studies have compared the shared neural components in the processing of semantic meaning in language and music by studying the double dissociation between memory-based and rule-based knowledge (22,24). Using familiar and unfamiliar melodies modified with unexpected in-key or out-of-key notes, researchers manipulated memory-based and rule-based knowledge, respectively. A double dissociation occurs where memory violations elicit N400 components, while rule violations elicit early negativities with a shorter latency, suggesting an extension of the rule/memory dissociation in language to music (22). Although research shows preserved musical semantic memory in adults with AD, it remains unknown whether unexpected memory violations in familiar musical stimuli would elicit such an N400 component response in this population.

Summarizing, there is compelling evidence of preserved musical semantic memory in patients with AD, suggesting that music-based interventions may contribute to delaying memory decline. Still, there is a lack of rigorous studies examining whether musical semantic memory for newly learned songs can be developed in patients with AD, and, if so, whether this ability is linked to general cognitive function. The overall aim of this clinical trial is to address this gap in research by exploring the connection between musical semantic memory and cognitive function, and by this follow-up on results from a recent case study showing memory for newly learned music despite clinical deterioration (17).

Based on outcome measures from observations of mood and behaviour, cognitive test performance, and EEG examinations, the present study aims to answer the following research questions:

1. Can an individual singing-based intervention improve AD patients’ musical semantic memory?
2. Does improvement in AD patients’ musical semantic memory relate to improvement of cognitive functioning?
3. Are improvements in musical semantic memory and cognitive functioning mediated by mood?

## OBJECTIVES

The primary objective of this study is to examine effects of intensive versus minimal individual musical training based on singing novel songs on memory for music and general cognitive functioning in non-musician adults with AD.

Secondary objectives are as follows:

- To determine the number of repetitions needed for independent recalling (total or partial) of new songs.
- To determine the quantity and quality of cues needed for such independent recalling.
- To determine EEG markers (e.g. N400) related to the sense of familiarity for new songs.
- To examine if intensive individual training improves memory for music performance.
- To examine if effects on cognition and memory for music are mediated by mood, and if effects on cognition are mediated by mood and memory for music (Fig. 1).

**Figure 1.**
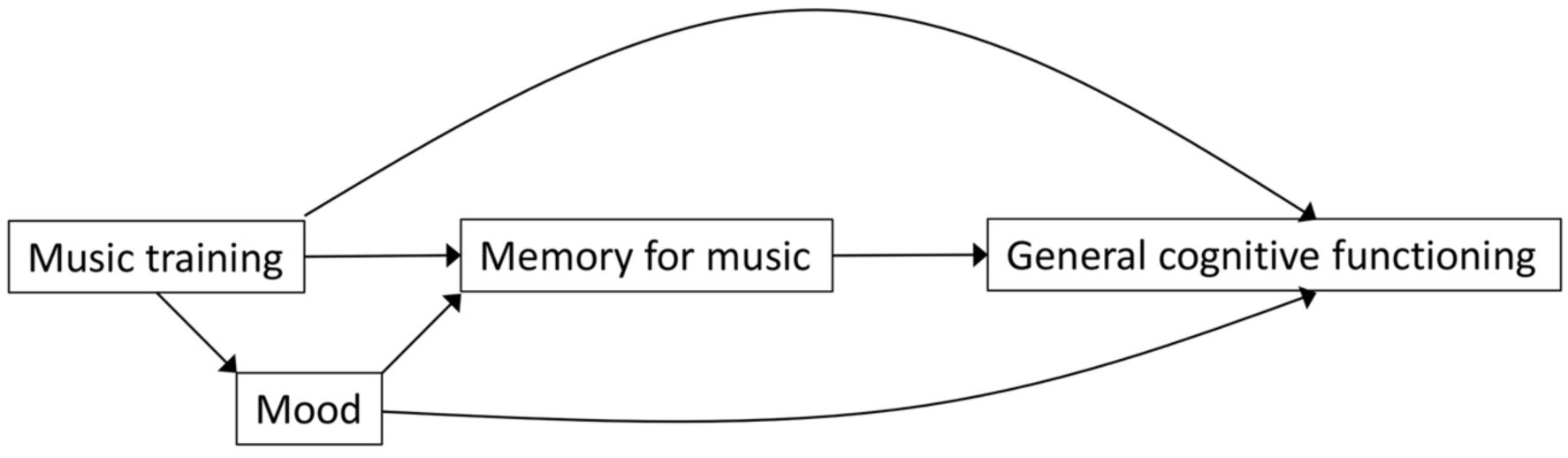
Path diagram of hypothesized intervention mechanisms and effects.

We hypothesize that participation in an intensive, individual singing-based intervention will improve the participants’ musical semantic memory, which in turn will be associated with improved cognitive function. This effect may be mediated by positive mood (Fig. 1). Furthermore, if participants with AD show an N400 component in response to unexpected in-key violation of a newly learned melody following the proposed intervention, it would indicate that musical semantic memory – as indexed through EEG measures – can be improved.

Findings gained from this study may have implications for evidence-based treatment planning and for designing opportune interventions aiming at reducing cognitive decline and improving the quality of life of people with AD. In line with the Global Action Plan on the Public Health Response to Dementia (25), the present study contributes by focusing on mechanisms of memory for music, particularly in an AD population, and by investigating whether musical interventions have any impact on overall cognitive functioning. Additionally, the findings could be adapted and replicated in other parts of the world to design culturally sensitive approaches for this population.

## METHODS

### Trial design and procedures

M4M is a prospective, experimental, crossover, before-and-after, international, assessor-blinded randomized controlled trial (RCT; Fig. 2). The main advantage of a crossover design compared to a simpler parallel design is to obtain the same statistical power with a reduced sample size, because effects can be compared within as well as between participants, thus accounting for inter-individual variation. There may be carry-over effects of the initial treatment (26), but they can be accounted for statistically through additional assessments before the second treatment.

**Figure 2.**
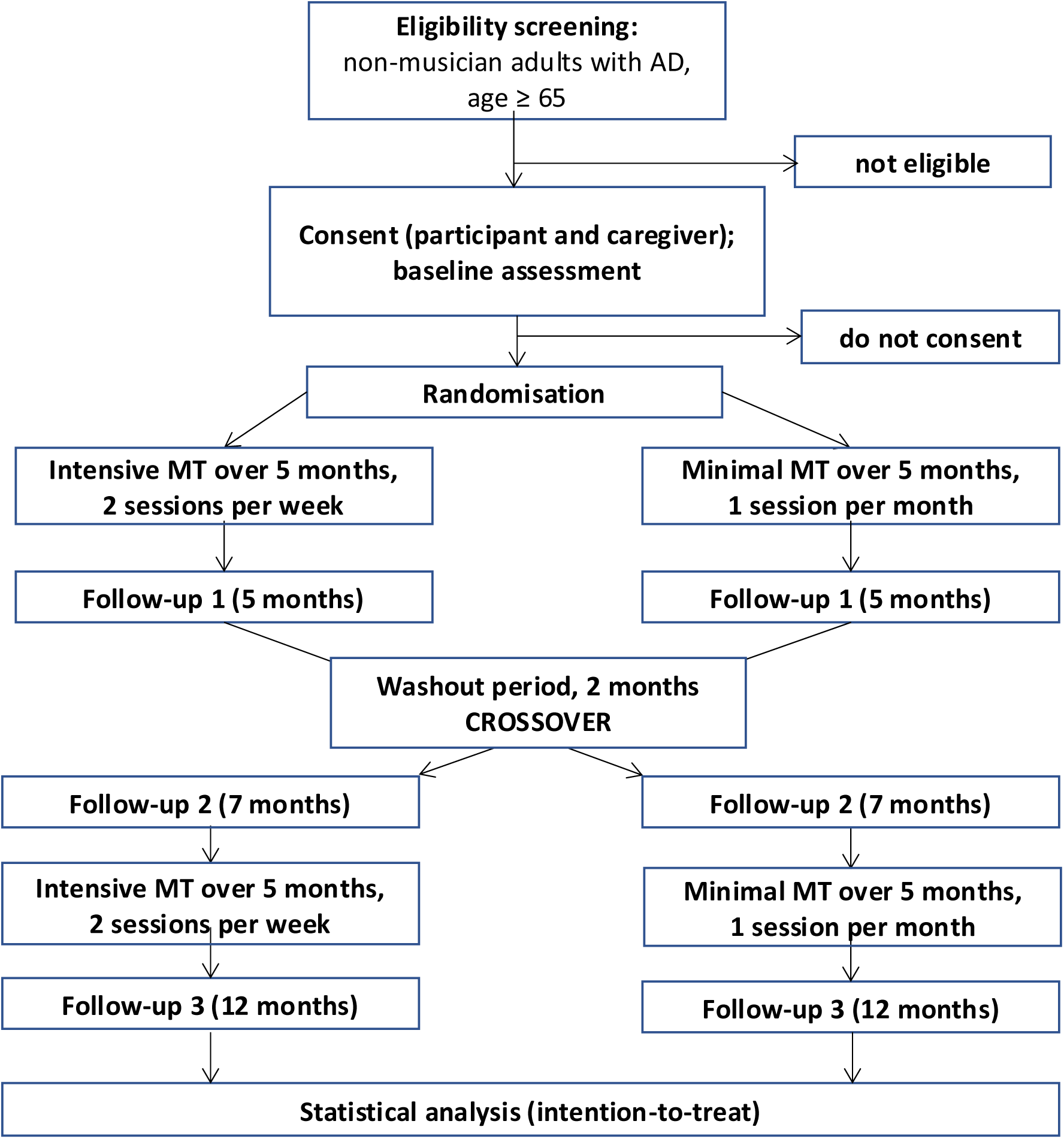
CONSORT flow diagram depicting the design. *Note.* AD, Alzheimer’s Disease; MT, musical training.

The interventions are based on previously described and tested models used with musician and non-musician adults with AD (15–17,27,28). In this context, the proposed approach builds on emerging knowledge and consensus related to the favourable outcomes secondary to the use of live music with this population. Eligible trial participants who have given consent to participate will be randomly allocated to receive musical training, either in a sequence of intensive followed by minimal intervention or vice versa with a washout period of 2 months in between the two phases. Intensive phase includes 2 sessions per week, minimal phase 1 session per month (Fig. 2 and 3).

**Figure 3.**
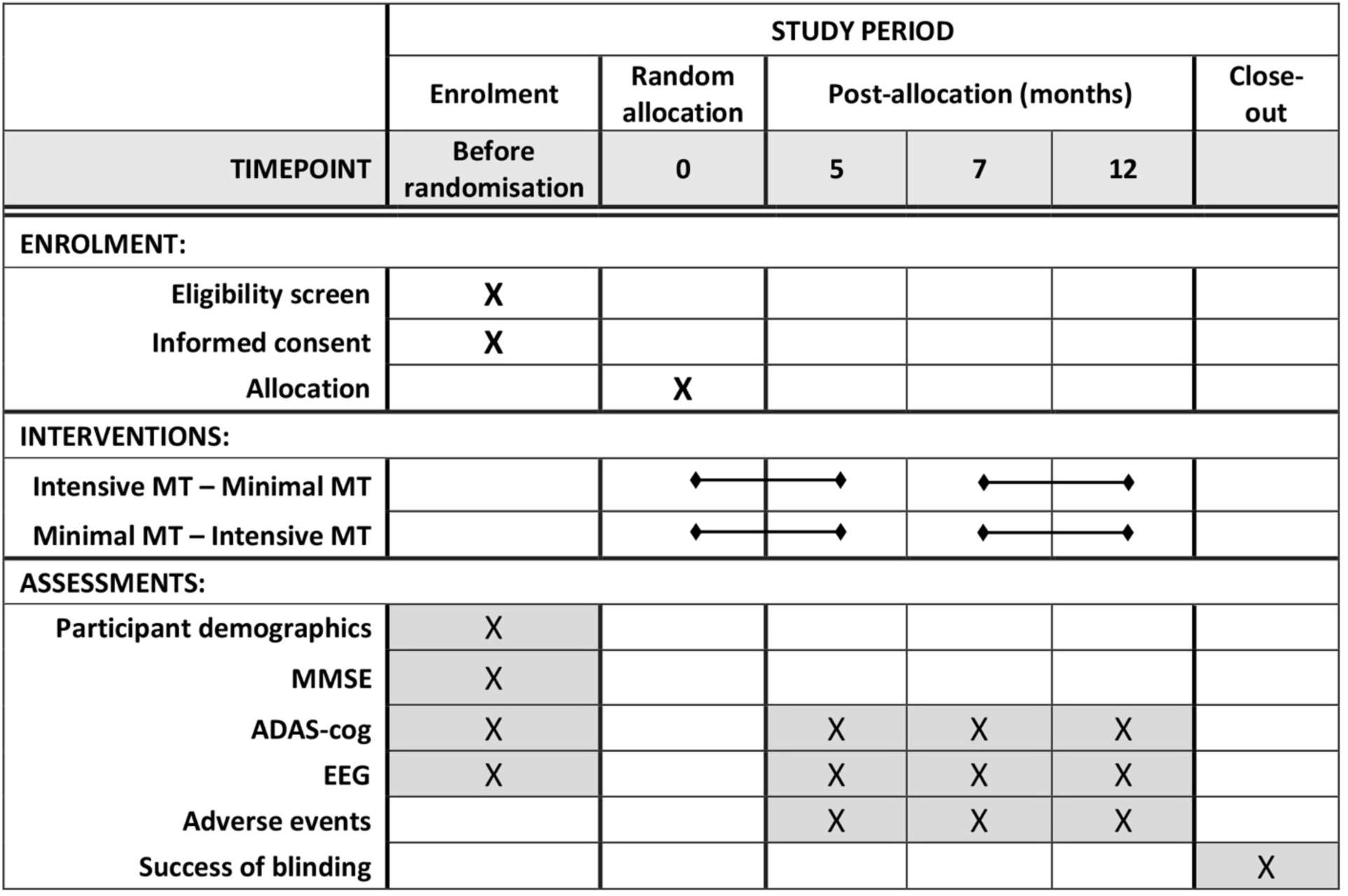
SPIRIT diagram depicting the data collection schedule. *Note.* ADAS-cog – Alzheimer’s Disease Assessment Scale – Cognitive Subscale; EEG – electroencephalogram; MMSE – Mini-Mental State Examination; MT – musical training.

Randomization will be conducted centrally at NORCE, concealed from site investigators, using block randomization with randomly varying block size, with separate lists for each site. Once a decision on inclusion has been made, informed consent obtained, and baseline data collected, the randomization result will be revealed to the instructors through an online system. Participants and instructors cannot be blinded due to the nature of the intervention. External evaluators will remain blinded after randomization; success of blinding will be verified at the end of each participant’s participation. Treatment fidelity (adherence and competence) will be evaluated by external raters using video recordings. This study protocol is designed and reported in accordance with the SPIRIT 2013 statement. Ethical approval has been obtained for Norway (Regional Committee for Medical Research Ethics South East Norway, 09 September 2024, reference number 759936) and will be obtained from the responsible ethics committees in each country prior to starting recruitment. The trial is registered in Clinicaltrials.gov, NCT06611878.

### Participants

We aim to recruit 113 non-musician adults with a diagnosis of probable or definite AD. To achieve the target sample size, it is necessary to recruit participants from multiple sites. Sites are located in Europe and South America (confirmed: Buenos Aires, Argentina; Bergen, Oslo, Kinn, Norway; pending funding: Vienna, Austria; see trial registration for updated list).

#### Eligibility criteria and enrolment

Participants eligible for the trial will be of any gender and ethnicity/nationality; aged ≥ 65 years; non-musician; have a documented diagnosis of probable or definite AD; and be home-dwelling in the vicinity of a study site. The research team will conduct the screening of the participants based on available results from neurological and neuropsychological examinations that support a profile compatible with AD, clinical history and musical engagement/history profile. Individuals with a confirmed non-AD dementia type (e.g. vascular, frontotemporal, Lewy body, mixed or pseudo-dementia), suffering from other known neurological disorders (e.g. Parkinson’s disease, multiple sclerosis, stroke), with known severe mental illness (e.g. current major depression, bipolar disorder, major anxiety, schizophrenia), or with known severe hearing loss that is not compensated by hearing aids will be excluded. For the purposes of this study, we consider as non-musicians those who have no history as professional musician (see Table 1 for history of revisions of this criterion). Participants should live within reasonable distance of a study site (e.g. within ≈1 hour driving distance), understand the language(s) used at the study site, and expect to be available for 1 year from enrolment. They may be under pharmacological treatment for AD and other diseases; such treatment should be stable at least 8 weeks prior to inclusion and will be recorded. Study participation will be based on informed consent from the participant. Enrolment will be voluntary and in response to a public recruitment call via media and advertisement flyers in day activity centres, hospital memory clinics, and assisted living organizations.

**Table 1.** History of protocol amendments.

| Date | Amendment number | Version name | Amendment | Rationale |
| --- | --- | --- | --- | --- |
| 2022 | - | Version 0 (original proposed by M.L.) | - | - |
| February 2023 | - | Version 1 (original submitted to RCN; accepted for funding by RCN in 2023; and submitted to REK for ethics approval in 2024) | (1) Design changed from parallel to crossover; (2) primary outcome changed to general cognition; (3) ERP stimuli specified further. | (1) Improved power; (2) direct patient relevance in line with funding call; (3) improved clarity. |
| June 2024 | 1 | Version 2 (this version; submitted for publication) | No MMSE cutoff. | MMSE scores depend on gender, age, and education level. Clinical criteria are more meaningful. |
| June 2024 | 2 | Version 2 | Clarified definition of “non-musician”: removed “no intensive training in childhood or youth up to 20 years of age; $\leq 2$ years of musical practice/training in adulthood”; retained only “no history as professional musician”. | Original additional criteria were unclear and did not work well across countries due to different education systems. |
| August 2024 | 3 | Version 2 | Recruit only participants with ability to provide informed consent. | Original was questioned by Norwegian ethics board. Number without ability to consent expected to be very low among otherwise eligible participants. |
| August 2024 | 4 | Version 2 | Definition of regions of interest (ROIs) for ERPs: removed specific electrode names in favour of more general description (6 ROIs, 2x3 grid; exact time window and ROI to be adjusted). | Electrode names may not be applicable to different EEG systems used across sites. |
| August 2024 | - | Version 2 | ERP stimuli and other design aspects specified further (e.g. only in-key violations); descriptions improved. | Further clarification of procedures. |

#### Baseline data

Demographics data, performance on the Mini-Mental State Examination (MMSE) (29), and baseline values of outcomes (described below) will be collected before random allocation. Participants’ history of musical training and general education will be recorded. The Music Engagement Questionnaire (30) will be completed by participants and/or caregivers, to provide insight into participants’ everyday life engagement with music, which may relate to musical semantic memory (31). In addition, we will collect information about the language(s) spoken by the participants, the age at which each was acquired, and the proficiency levels they had attained before starting to suffer from cognitive decline. Where necessary, this information will be collected with the help of their next of kin. Several outcomes will also be collected at baseline (see Outcomes and Fig. 3).

### Interventions

Interventions will be conducted by an instructor with adequate qualification and experience in music teaching and dementia care. Instructors will ideally have a music therapy degree, but music educators with relevant experience and training, supervised by music therapists, may also be able to conduct singing interventions for dementia (7). Interventions will entail individual musical training based on singing to learn novel songs. These songs will be composed for the purpose of this study and will take into consideration adequate musical complexity, length of verses and chorus, and high-frequency words for the lyrics. The structure of the complete lyrics of the song will have the following sequence (similar to popular music): first verse; second verse; chorus; repetition of the first two verses; chorus; repeat chorus; final cadence.

Participants will be randomly assigned to one of the two sequences: intensive intervention phase (two sessions per week) followed by minimal intervention phase (one session per month) or vice versa (Fig. 2). They will receive individual musical training based on singing during both phases, lasting five months each, with two months of no intervention in between. During both intervention phases, participants will learn and focus the singing practice on one novel song per month, that means, a total of five novel songs will be offered in each phase. The order of the songs will be randomised across the 10 months of intervention.

The instructor will use live music by singing with the support of a harmonic instrument such as piano, keyboard, guitar or accordion to deliver the musical experiences. At all times, the instructor must provide appropriate and individualised support encouraging the participant to engage in the tasks without forcing him/her to do so. It is crucial that the instructor establishes an empathic relationship with the participant for the musical experience to be pleasant, rewarding, emotionally meaningful and conducive to learning. Each session will have a duration of 40 to 45 minutes and will be conducted according to a sequence of steps (19) as follows:

*Step 1) Baseline mood observation:* The instructor will show a set of icons (Supplementary Fig. S1) to the participant to self-report his/her mood and will note the response on the Scoring Session Instructor Form (SSIF, Supplementary Form F1). If this is not possible, the instructor will complete it based on observed behavioural/gestural/verbal responses.

*Step 2) Warm up:* The instructor will offer a 5-minute warm-up which includes breathing exercises and vocalizations. Five breathing exercises will be performed at the beginning of the lessons to practice breathing, air dosing, as well as to promote relaxation and prepare the participant for singing. After this, vocalizations will be performed. They must be offered in a pitch range matched to the participant’s vocal range. It will entail a sequence of two vocalizations such as: a) major thirds by ascending and descending steps, b) perfect fifths by ascending and descending steps. The rhythmic pattern suggested is a dotted eighth note with a sixteenth note (Supplementary Fig. S2), sung in a playful manner to promote positive mood.

*Step 3) Teaching the song*: First, the instructor will introduce the new song by singing it through once. A key which is comfortable for the participant will be used in all cases. The instructor will then complete a 5-item Likert scale to describe the participant’s response (liking or disliking) to the new song based on observed behavioural/gestural/verbal responses (5-point Likert scale; Supplementary Form F1). The instructor will then teach the song. By applying their own judgement, the instructor will decide which didactic resources and strategies are best to teach the song based on the participant’s preferred learning style and performance. The instructor might choose one of the following teaching strategies: a) repeatedly singing the entire song together with the participant having the printed lyrics as support; b) repeatedly singing together with the participant one line at a time until completion of the chorus first and then the verses of the song, having the printed lyrics as support; c) without having printed lyrics, the participant will learn by imitation one line at a time or the entire song. If choosing the option of teaching one line at a time during the *intensive* intervention and one session is not enough to work on the entire song, the verses and the chorus will be learned in two different consecutive sessions. Each line should be repeated between 4 and 6 times to provide opportunities to consolidate learning. If choosing the option of teaching one line at a time during *minimal* intervention, the instructor will begin teaching the chorus first, and if there is sufficient remaining time will teach the verses. The instructor will offer the type and number of cues that the participant needs to successfully perform the task of learning and recalling the song. The instructor will use their own judgment to gradually decrease or increase cues and support depending upon participant needs to successfully complete the singing task.

*Step 4) Singing the entire chorus:* The participant will be invited to sing the entire chorus. The instructor will provide the type and amount of assistance/support as needed by the participant to successfully perform the singing task. This step should be repeated eight times.

*Step 5) Singing chorus in context:* The participant will be invited to perform the entire song together with the instructor. The instructor will sing the verses solo, and the participant will sing solo the chorus. The instructor will provide support of the printed lyrics of the entire song. The participant may be inclined to join singing during the verses as well; the instructor will give space for him/her to do so. A brief musical instrumental introduction will be offered to establish the musical structure and character of the song and to orient the participant in tonality, meter, character and tempo of the song. The instructor will provide the type and amount of assistance/support as needed by the participant to successfully perform the singing task. This step should be repeated at least twice. At each song ending, instructor and participant may engage in a brief verbal exchange about the singing experience or the content of the song, which will work as a brief pause, release and distractor before repeating the full song a second time.

*Step 6) Practicing previously learned new song:* Starting in month two of the first musical training phase, the instructor will invite the participant to rehearse a novel song learned in a previous month (Table 2). The intervention will be conducted as described in *Step 5*.

**Table 2.** Combination of Songs for Rehearsal (example with minimal intervention first)

**a) Minimal intervention phase**
| **task** | **month 1** | **month 2** | **month 3** | **month 4** | **month 5** |
| --- | --- | --- | --- | --- | --- |
| new song | song 1 | song 2 | song 3 | song 4 | song 5 |
| rehearse | - | song 1 | song 2 | song 3 | song 4 |

**b) Intensive intervention phase**
| **week** | **task** | **month 8** | **month 9** | **month 10** | **month 11** | **month 12** |
| --- | --- | --- | --- | --- | --- | --- |
| 1 | new song | song 6 | song 7 | song 8 | song 9 | song 10 |
|  | rehearse | - | song 6 | song 7 | song 8 | song 9 |
| 2 | new song | song 6 | song 7 | song 8 | song 9 | song 10 |
|  | rehearse | - | - | song 6 | song 6 or 7 | song 6, 7, or 8 |
| 3 | new song | song 6 | song 7 | song 8 | song 9 | song 10 |
|  | rehearse | - | song 6 | song 7 | song 8 | song 9 |
| 4 | new song | song 6 | song 7 | song 8 | song 9 | song 10 |
|  | rehearse | - | - | song 6 | Song 6 or 7 | song 6, 7, or 8 |

*Note.* This is an example with minimal intervention first. The order is reversed for those randomised to intensive first.

*Step 7) Session closure:* The purpose is to relax and bring the session to closure. If the participant is available and willing to do so, he/she will choose a familiar favourite song and will sing it once together with the instructor. The instructor will provide support of printed lyrics if needed. The instructor will accompany the participant with the harmonic instrument (piano, keyboard, guitar or accordion) for the participant to accomplish this task. A comfortable tonality for the participant will be used in all cases. The instructor will decide moments of solo and tutti depending on the participant and will provide a musical context by sensitively responding to the musical and communicative expressions of the participant.

*Step 8) Final mood observation:* The instructor will again use a set of icons (Supplementary Fig. S1) and note the response or observed behaviour (Supplementary Form F1) to describe the participant’s final mood as in *Step 1*.

*Step 9) Session rating:* After the session, the instructor will complete the remainder of the SSIF (Supplementary Form F1) to record what happened in the session, including the participant’s responses and performance, title of the intervention song, title of favourite song, duration of the session, the degree of independent evocation and sense of familiarity (SoF) (15,34). The participant’s independent recall will be rated as follows: a) needs full support; b) needs intermittent support to complete each line; c) needs support in one syllable/word per line; d) fully independent. To record independent evocation, the instructor will indicate with an “X” when the first independent recall occurs during the session, that means, they will mark only one cell in the form depending on type of recall and in which repetition it occurred. If the degree of independent recall changes over the same session, the instructor will also indicate with an “X” the maximum degree of independent recall where it applies. Otherwise, if remaining stable, “no change” will be recorded on the SSIF.

Ratings of mood and SoF will also be analysed as outcomes (see Behavioural outcomes).

#### Assessment of intervention fidelity

External evaluators blinded to intervention type and session number will rate selected sessions at specific points of the training (last session of each month and when the instructor’s SoF rating changes). To that end, sessions will be video recorded to allow external blinded evaluators to audit the session to determine adherence to the intervention manual and competence by completing the Intervention Fidelity Form (Supplementary Form F2).

### Outcomes

The main outcomes, including neuropsychological assessments (33) and EEG, will be measured before and after each musical training phase (Fig. 3). Behavioural assessments will be conducted by the instructors in every session and by external evaluators in selected sessions at specific points of the training (last session of each month and when the instructor’s SoF rating changes).

#### Behavioural outcomes

*Mood* (Step 1 and 9 in Interventions) at the beginning and end of each session will be measured on 5-point Likert scales.

*Sense of familiarity* with the current song will be assessed using the SoF scale (15,34), a 6-point Likert scale. It will be completed by the instructor at the end of each session (Supplementary Form F1) and by the external evaluator after selected sessions (Supplementary Form F3). For this study, a score of 4 or above will be considered as the threshold to determine familiarity-based recognition.

*Performance* as observed by an external evaluator based on video recordings will be analysed in selected sessions using the Participant Performance Cue Indicator Form (PPCIF, Supplementary Form F3). It yields a score for chorus solo (range 8-32, where 32 is best) and for chorus in context (2-8, where 8 is best).

#### Neuropsychological outcomes

Before and after each musical training phase, a neuropsychological assessment (ADAS-cog) (33) will be administered by a trained psychometrician blinded to the intervention allocation. The scale consists of items to assess the following domains: language; memory; praxis; and orientation. The standard ADAS-cog includes the following 11 subtests, with a total scoring range from 0 (no impairment) to 70 (most severe impairments): word recall (0-10); commands (0-5); naming (0-5); constructional praxis (0-5); ideational praxis (0-5); orientation (0-8); word recognition (0-12); remembering word recognition test instructions (0-5); spoken language ability (0-5); comprehension of spoken language (0-5); word-finding difficulty (0-5). The ADAS-cog score is based on the number of errors made within each subtest. Two versions will be included: one administrated at baseline and at the 7-month assessment; another at the 5-month and 12-month assessments. In a r-test situation, the minimum clinically important difference (MCID) is 3 (34). The ADAS-cog takes 30-35 minutes to administer. The total score of the ADAS-cog at the end of each intervention will be used as the *primary clinical outcome measure*, and the scores on the eleven sub-domains as secondary clinical outcomes. We will aim to conduct neuropsychological assessments within seven days before the first and after the last musical training session of each intervention phase.

#### Neurophysiological outcomes

At the beginning and end of each intervention phase (Fig. 3), brain and behavioural response (SoF) during music recognition tasks will be recorded and analysed as outcome measures. We seek to determine the N400 component related to the in-key violations in songs learned during the intervention period and the potential correspondence with the behavioural observations. If participants do develop musical semantic memory through our intervention, we expect them to also show an N400-like component to in-key violations in familiarized melodies, i.e., those melodies that are introduced and learned during our intervention, but only after the intervention. If an N400 component is detectable during these in-key violations of a newly learned song, it will indicate memory of the new song even if the participant is unable to adhere to a behavioural task or display an observable sense of familiarity.

##### EEG protocol

Each site will collect EEG with at least 32 channels based on the 10-20 system. Preferably active EEG systems will be used which are less sensitive to interference from other sources, which is especially important when recordings are made outside shielded rooms (e.g. at participants’ homes). EEG recordings will start with a resting state measurement to familiarize participants with the situation of the EEG data collection. For the purposes of our study, a previously used paradigm (22) is adapted to have in-key violations in familiarized/to-be-familiarized and unfamiliarized melodies only to assess reactions to memory-based knowledge. No out-of-key violations will be introduced. The in-key violations will be introduced at multiple places throughout each melody, which will consist of at least 32 bars. No violations will be introduced in the first four bars, to allow participants sufficient time to recognize the melody. A total of 10 melodies will be played for each participant, of which half are learned in an intervention period (familiarized/to be familiarized). In each of the melodies we will include at least 8 in-key violations. ERPs in response to these in-key violations will be compared to ERPs in response to unchanged notes in structurally similar places in each melody. After each melody, applause will sound. Participants will be instructed to indicate via a button when they hear the applause. We estimate that the paradigm will last about 20 minutes. Half of the melodies will be and remain novel or unfamiliarized, that is, they will not be introduced during the intervention. The other half of the melodies will be novel at the first EEG assessment but be introduced during the intervention and thus be considered familiarized at the second EEG assessment (familiarized/to be familiarized). At least forty in-key violations will appear in novel melodies, and at least forty in-key violations will appear in familiarized melodies. EEG assessments will be conducted within seven days before the first and after the last musical training session of each intervention phase. If memory for music is gained through the intervention, we expect more pronounced N400 amplitudes to in-key violations in familiar melodies but not to in-key violations in unfamiliar melodies after the intervention. Thus, we expect an interaction effect of time (before vs. after intervention period) and melody type (familiarized/to be familiarized vs. unfamiliarized) on N400 amplitude variation.

##### EEG data analysis

EEG raw data will be converted to MATLAB-readable files and pre-processed within EEGLAB according to recommended processing pipelines. General EEG preprocessing steps will include bandpass filtering, artifact rejection, including the removal of noisy channels and time segments, and eye movement components identified via independent component analysis (ICA, (35)), before data epochs will be extracted for trials with or without memory violations for familiarized and unfamiliarized melodies. We will examine differences in mean amplitude in response to in-key violations and unchanged notes at six regions of interest (ROIs) based on previous research in a two-by-three grid with two levels of anterior/posterior and three levels of laterality (left, mid, right) (22,24). The exact time window and ROI studied may be adjusted given the potential for an altered latency and amplitude of the N400 component in the participants (20) and the use of different EEG systems between sites using cluster-based permutation tests.

#### Adverse events

Although adverse events (AEs) of music interventions are rare (36), AE assessment is an important part of the safety aspect in all RCTs. We will ask participants or relatives at each assessment time point about the occurrence and a description of any serious or non-serious AEs. Potentially study-related serious AEs, as well as unblinded frequency counts of any AEs, will be shared and discussed with the Data Monitoring and Safety Committee (DSMC) through the trial statistician.

### Sample size and power, data monitoring, statistical analysis

#### Sample size and power

Although cognitive outcomes have been of secondary interest in music interventions in dementia so far (6) there is some research to suggest a plausible effect size. Our systematic review found a mean d=0.29 (95% CI 0.02 to 0.57) of active music therapy on global cognition, based on 3 RCTs (total 209 participants); in contrast, no effects were found for music listening (37). A study of group-based singing found similar effects (38). Analyses in the Cochrane review did not distinguish between active and listening interventions (7). The ADAS-cog was only used in one included study (7), although it is more sensitive than the MMSE (39) and has a defined MCID (34). With a conservative assumption of SD=10 (a large sample had SD=6.42 (39)), the MCID of 3 corresponds to an effect size of d=0.30, similar to our previous review (37). Further assuming a correlation r=0.5 between interventions, attrition ≤ 20%, and aiming for 80% power with a two-sided 5% significance level leads to a required number of 113 participants to randomize (90 after drop-out). Similar power would be reached for slightly smaller d and higher r or vice versa; higher power if SD is smaller. In contrast, a parallel trial would require 440 participants to achieve the same power for the same effect size. Recruitment and follow-up rates as well as data quality will be monitored and discussed with the DSMC to ensure target sample size and power.

#### Statistical analysis

Sociodemographic and diagnostic features will be analysed via descriptive methods (mean [SD], range, n [%]). Outcomes will be analysed on an intention-to-treat basis (analysing all participants as randomised, regardless of actual participation), which provides a conservative estimate of effects, and additionally on a per-protocol basis (analysing participants according to their actual participation in the trainings). We estimate that participants need to receive around 90% of the sessions during intensive musical training intervention for learning effects to occur. Multiple imputation will be used if attrition is higher than expected. Continuous outcomes will, following a graphical examination of normality, compare the post-test difference between interventions within each participant, adjusted by the difference in pre-tests within each participant in a hierarchical ANCOVA model with participant and site as random effects. The ANCOVA model ensures optimal use of baseline measures to improve precision and avoids bias from randomly occurring baseline imbalance (40). The model can be written as *(Y_Ti_ – Y_Ci_) = β_T_ + γ(X_Ti_ – X_Ci_)*, where *Y* is the post-test, *X* is the pre-test, *T* is the treatment, *C* is the control condition, *γ* represents the influence of the pre-test, and the intercept *β_T_* represents the treatment effect (40). However, as this model does not address possible carry-over effects of the initial treatment (see Trial design and procedures), we will also include the randomisation (intervention sequence) in the model. Secondary analyses will include additional covariates/subgroups such as sex, age, AD stage, history of musical training, general education, music engagement, and interventionist’s background training (music therapy vs. music education). The role of mediators will be analysed with a series of regression models (as depicted by each arrow in Fig. 1). Data will be analysed using R (r-project.org).

## Discussion

As the most common form of dementia, AD is one of the major causes of disability and dependency among older people. A global action plan issued by the WHO aims to “improve the lives of people with dementia, their carers and families, while decreasing the impact of dementia on them as well as on communities and countries.” (25). Based on previous evidence from trials on music and dementia with promising effects on mood and behaviour problems, M4M focuses on cognitive outcomes in home-dwelling older adults with AD, thus aiming to prevent or decelerate further decline in their abilities. Using an intensive singing intervention to increase the ability to learn and recall new songs, this trial is in line with the goal for people with AD to experience greater mastery and to live active and meaningful lives with cultural activities adapted to individual interests and needs (25). The intervention is relevant for all genders and can be tailored to different cultural backgrounds. Providing individualized services to home-dwelling service users is in line with the aim of enabling people with AD to live with home care outside of care facilities as long as possible.

M4M will be the first study to rigorously examine musical semantic memory for newly learned songs in patients with AD receiving individual musical training. By combining behavioural and electroencephalographic (EEG) measures and relating them to performance on neuropsychological tests, we will further be able to explore how musical semantic memory may benefit cognitive functioning. Through EEG measurements, changes in brain processing in people with AD will be examined as a potentially more sensitive marker than behavioural and verbal responses to predict clinical outcome (Fig. 1). Our results may thus help move the field towards finding patient-tailored therapies (41).

Finding ways to alleviate dementia symptoms, understanding the mechanisms by which potential interventions work, and exploring the participant-specific factors that may contribute to intervention success are all crucial to cushion the enormous socioeconomic impact that dementia has. We expect to establish memory for music as an important mechanism and predictive marker of clinical improvement. In addition, we will also contribute to our fields’ understanding of the neural bases of musical semantic memory as well as the structure of musical semantic memory. M4M features interdisciplinary collaborations between neuropsychologists, musicologists, music therapists, music educators, trial methodologists, bioengineers, neuroscientists and biostatisticians. It will contribute to future practice development in dementia care, and to theory and research in the fields of music, health, aging, and neuroscience. Findings will be directly applicable to clinical practice and will have a valuable impact for individuals with AD, their family members, and practitioners working with them.

## Ethics

Approval for the first country has been received by the Regional Committees for Medical and Health Research Ethics, details see text.

## Dissemination

Dissemination of the findings will apply to local, national and international levels. Results will be published in specialised international journals. We will disseminate results through presentations at national and international conferences. In addition, dissemination, particularly to users, their families, user organisations, health services, and the general public will be achieved via media, educational brochures and/or popular press articles.

## Author Contributions

ML conceptualized and conceived the idea of the study, developed the original study design and protocol, and drafted, reviewed and edited the manuscript. CT provided scientific expertise to formulate the original study design and protocol. AXC, SK, DMP and EL contributed to the refinement of the neurophysiological outcomes. AJL contributed to the refinement of the neuropsychological outcome measures and AJL and RES with the presentation of clinical aspects of AD. CG and JA provided statistical expertise in clinical trial design and statistical analysis. AXC, CG and MG contributed to the funding acquisition. All authors reviewed and approved the final manuscript.

## Data Availability

Not applicable, as this is a study protocol.

## Acknowledgments

In Argentina, Jorge Juri and Carlos Canova have provided scientific advice on the original study design and helped to ensure the societal relevance of the study; Fundación Cuidados de Salud (Fernando Gril) have helped to ensure the societal relevance of the study. In Norway, Kinn Municipality (Jorunn Bakke Nydal); Oslo Municipality (locations: Myrer Omsorg+ and Pastor Fangens vei 22 Seniorhus; Elin Linløkken); the Norwegian Association for Music Therapy (Christine Wilhelmsen); and Oslo Dementia Association (Thale Waaler Eggen) have helped to ensure the societal relevance of the study. Narly Golestani has helped with additional measures and extensions to the study. Laura Fusar-Poli; Antoni Rodriguez-Fornells; and Teppo Särkämö have provided scientific advice on the study design.

## SUPPLEMENTARY MATERIAL – MEMORY FOR MUSIC PROTOCOL

**Supplementary Fig. S1.**
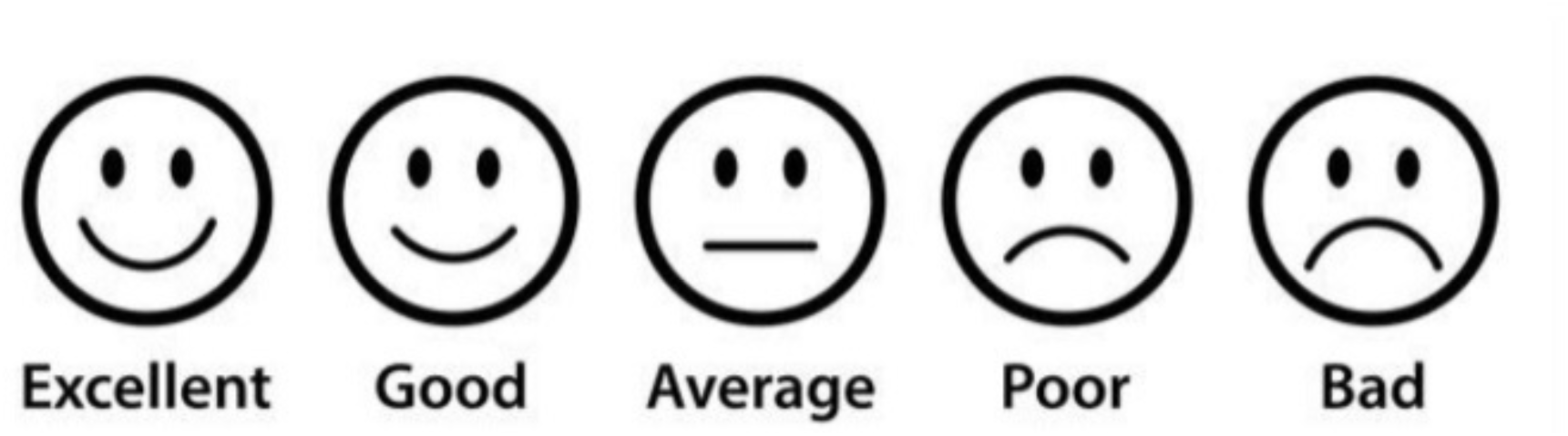
Likert Scale: Mood.

**Supplementary Fig. S2.**
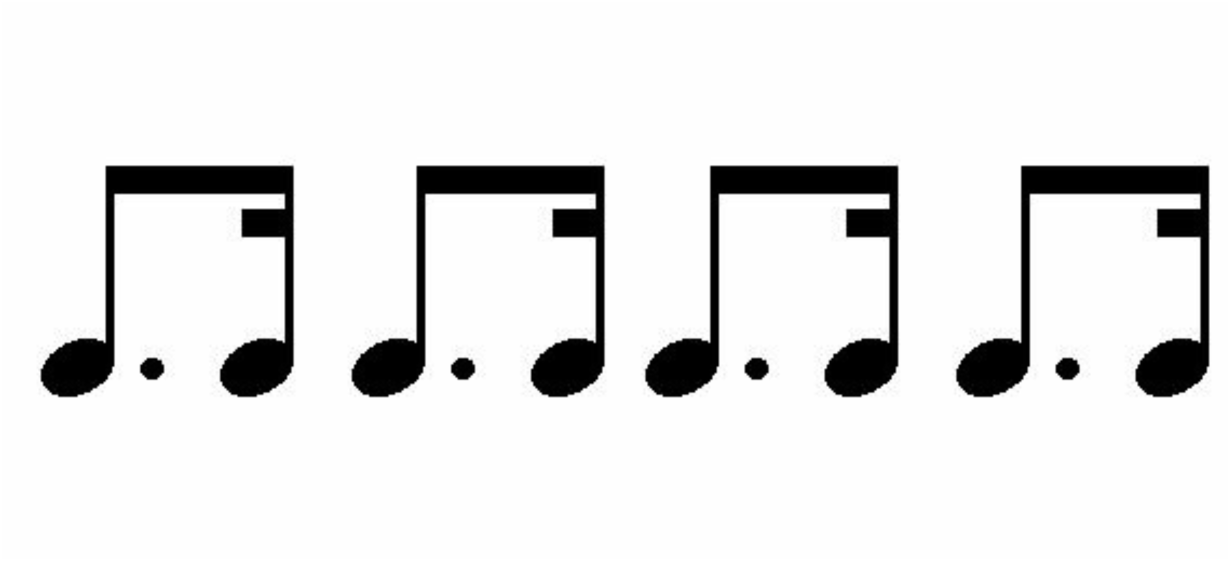
Pattern for vocalizations.

**Supplementary Form F1.**
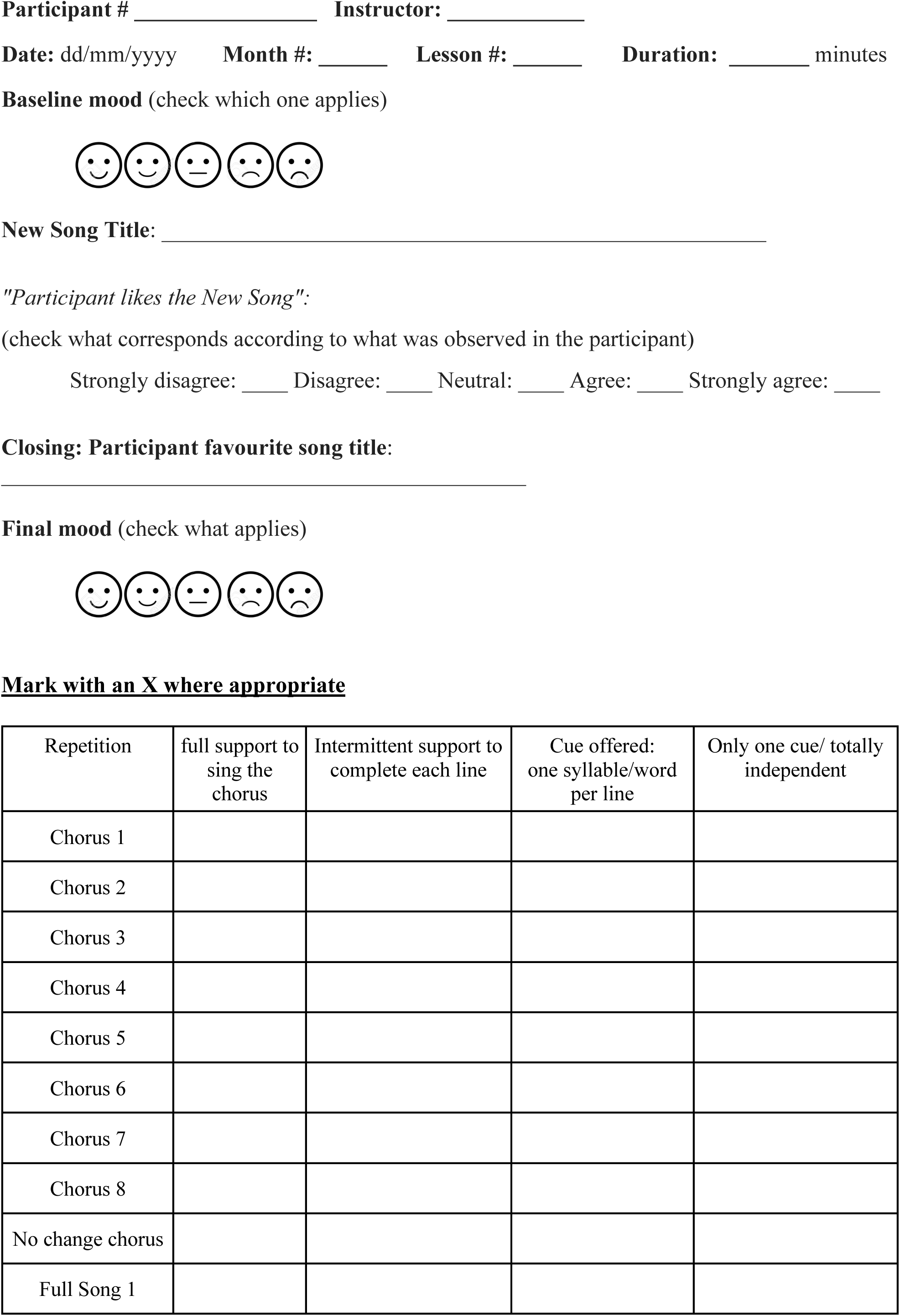
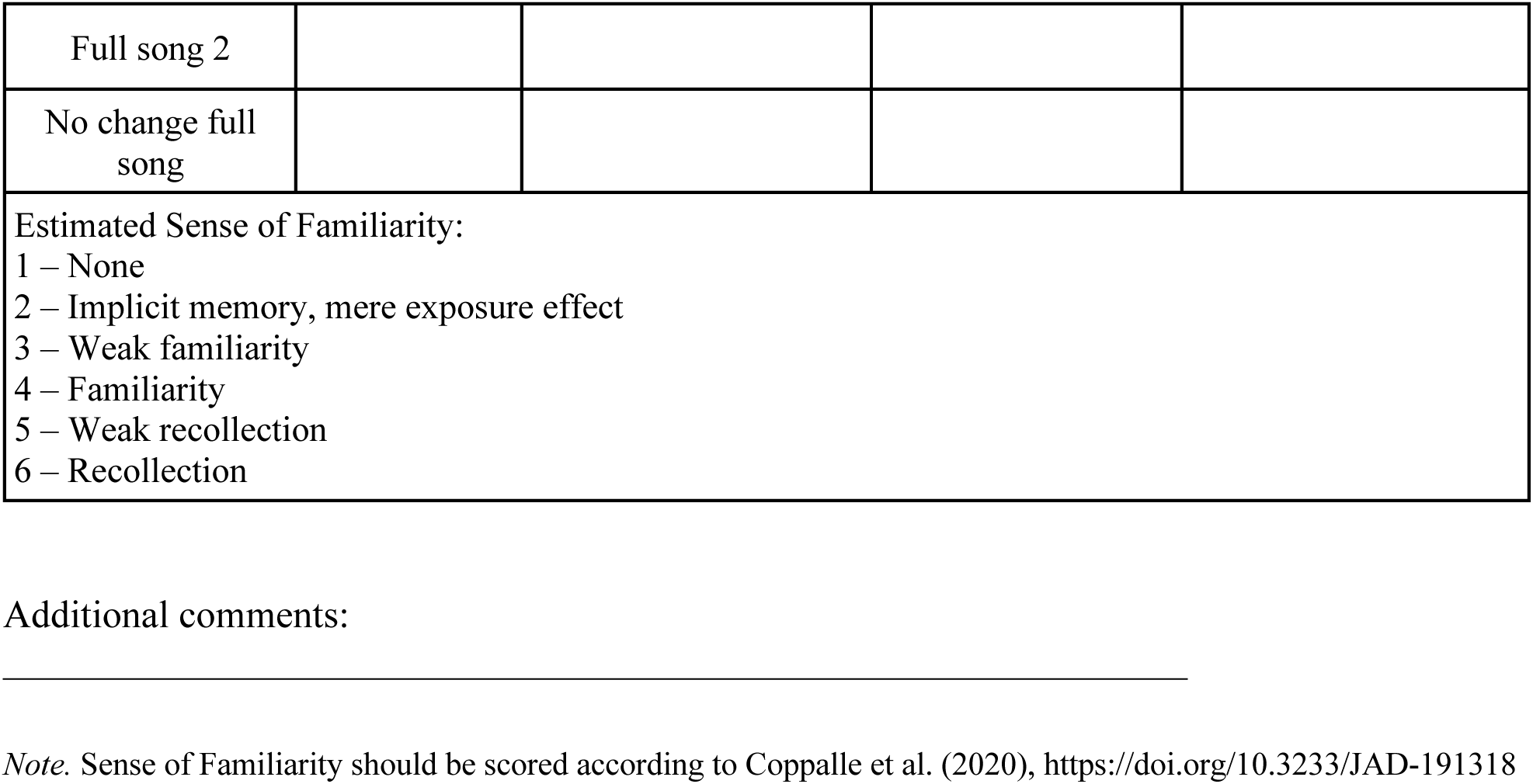
Scoring Session – Instructor Form (SSIF)

**Supplementary Form F2.**
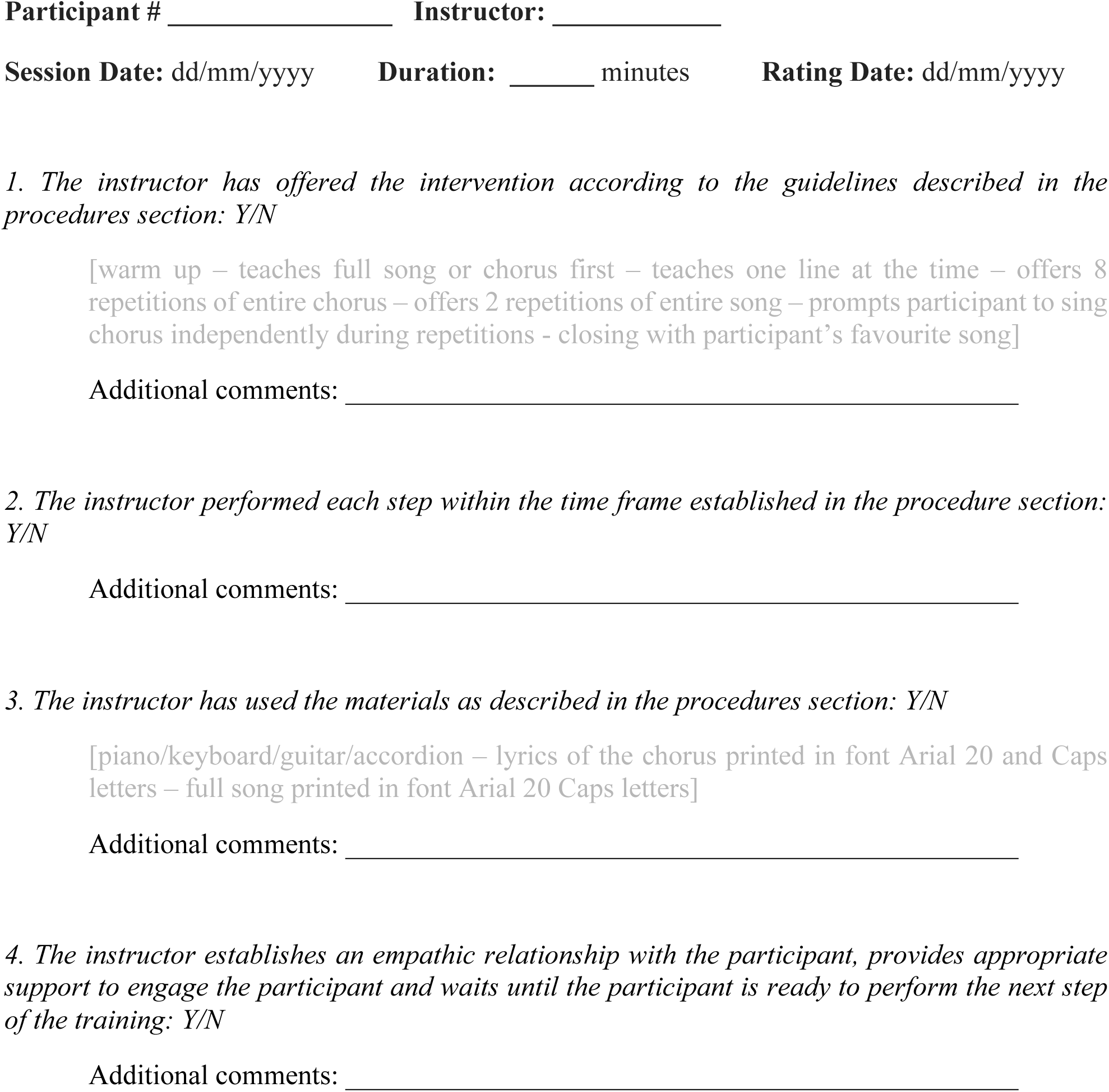
Intervention Fidelity Form.

**Supplementary Form F3.**
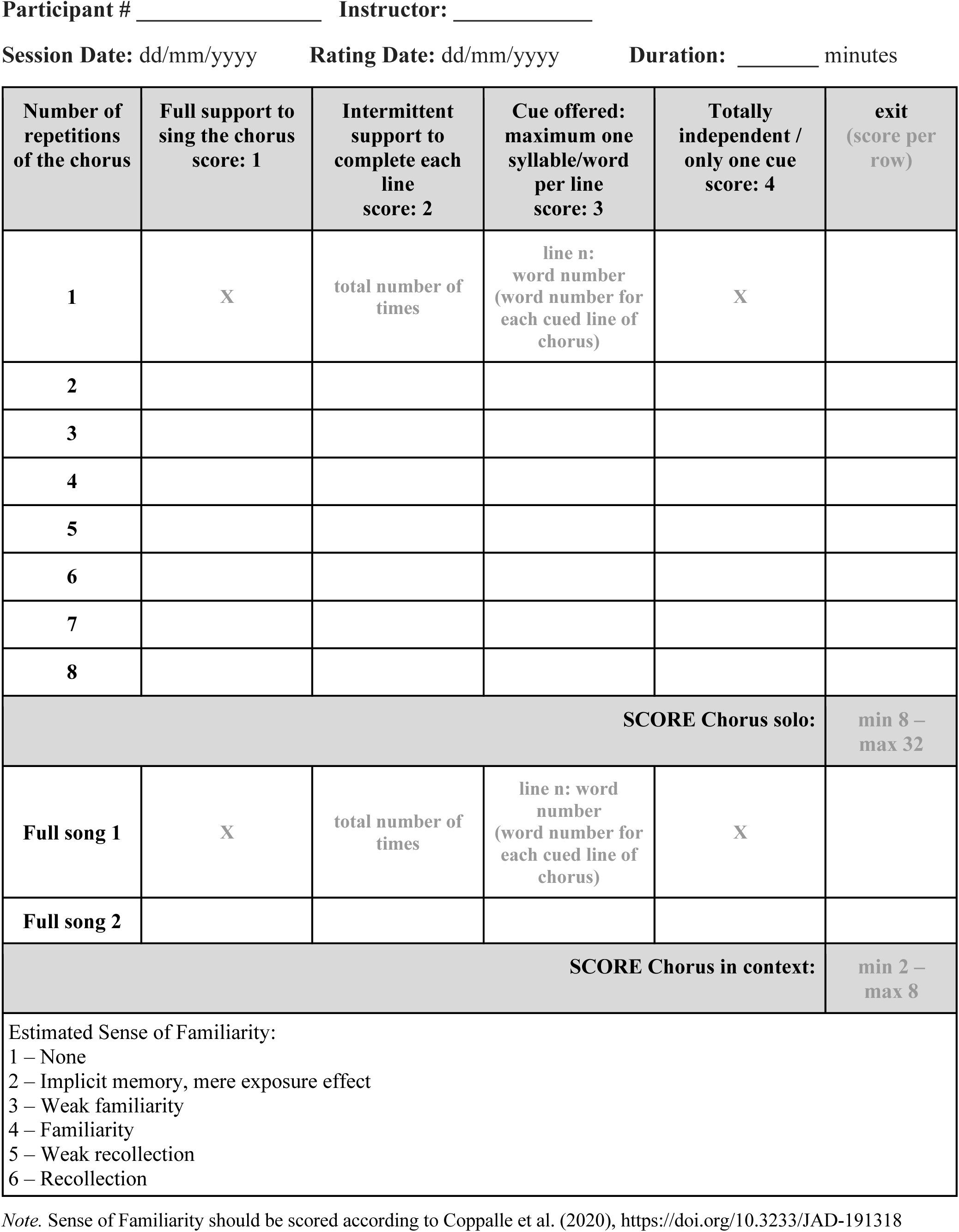
Participant Performance Cue Indicator Form (PPCIF) Used by external evaluator for scoring sessions.

## References

1. International Alzheimeŕs Disease. World Alzheimer Report 2019, Attitudes to dementia. Alzheimer’s Disease International: London. London; 2019.

2. Alzheimer’s Association. Alzheimer’s facts and figures report. Alzheimer’s Association. 2022;1.

3. Jahn H. Memory loss in Alzheimer’s disease. Dialogues Clin Neurosci. 2013;15(4):445– 54.

4. Cole JH, Franke K. Predicting age using n euroimaging: Innovative brain ageing biomarkers. Trends Neurosci [Internet]. 2017;40(12):681–90. Available from: 10.1016/j.tins.2017.10.001

5. Horvath A, Szucs A, Csukly G, Sakovics A, Stefanics G, Kamondi A. EEG and ERP biomarkers of Alzheimer’s disease: A critical review. Frontiers in Bioscience - Landmark. 2018;23(2):183–220.

6. van der Steen JT, Smaling HJA, van der Wouden JC, Bruinsma MS, Scholten RJPM, Vink AC. Music-based therapeutic interventions for people with dementia. Cochrane Database of Systematic Reviews. 2018;2018(7).

7. Baker FA, Lee Y eun C, Sousa TV, Stretton-smith PA, Tamplin J, Sveinsdottir V, et al. Clinical effectiveness of music interventions for dementia and depression in elderly care ( MIDDEL): Australian cohort of an international pragmatic cluster-randomised controlled trial. Lancet Healthy Longev [Internet]. 2022;3(3):e153–65. Available from: 10.1016/S2666-7568(22)00027-7

8. Mansens D, Deeg DJH, Comijs HC. The association between singing and/or playing a musical instrument and cognitive functions in older adults. Aging Ment Health [Internet]. 2018;22(8):964–71. Available from: 10.1080/13607863.2017.1328481

9. Crystal HA, Grober E, Masur D. Preservation of musical memory in Alzheimer’s disease. J Neurol Neurosurg Psychiatry. 1989;52(12):1415–6.

10. Cuddy LL, Duffin J. Music, memory, and Alzheimer’s disease: Is music recognition spared in dementia, and how can it be assessed? Med Hypotheses. 2005;64(2):229–35.

11. Cuddy LL, Sikka R, Vanstone A. Preservation of musical memory and engagement in healthy aging and Alzheimer’s disease. Ann N Y Acad Sci. 2015;1337(1):223–31.

12. Vanstone AD, Cuddy LL. Musical memory in alzheimer disease. Aging, Neuropsychology, and Cognition. 2010;17(1):108–28.

13. Groussard M, Chan TG, Coppalle R, Platel H. Preservation of musical memory throughout the progression of Alzheimer’s disease? Toward a reconciliation of theoretical, clinical, and neuroimaging evidence. Journal of Alzheimer’s Disease. 2019;68(3):857–83.

14. Ferreri L, Mas-Herrero E, Cardona G, Zatorre RJ, Antonijoan RM, Valle M, et al. Dopamine modulations of reward-driven music memory consolidation. Ann N Y Acad Sci. 2021;1502(1):85–98.

15. Coppalle R, Mauger C, Quernet S, Dewald A, Letortu O, Desgranges B, et al. New long-term encoding in severely amnesic Alzheimer’s disease patients revealed through repeated exposure to artistic items. Journal of Alzheimer’s Disease. 2020;76(4):1567–79.

16. Samson S, Dellacherie D, Platel H. Emotional power of music in patients with memory disorders: Clinical implications of cognitive neuroscience. Ann N Y Acad Sci. 2009;1169:245–55.

17. Lichtensztejn M. Memory for music and Alzheimer’s Disease. Case report. Salud, Ciencia y Tecnologia. 2022;2(2).

18. Lau EF, Phillips C, Poeppel D. A cortical network for semantics: (De)constructing the N400. Nat Rev Neurosci. 2008;9(12):920–33.

19. Federmeier KD, Kutas M. Aging in context: Age-related changes in context use during language comprehension. Psychophysiology. 2005;42(2):133–41.

20. Joyal M, Groleau C, Bouchard C, Wilson MA, Fecteau S. Semantic processing in healthy aging and Alzheimer’s disease: A systematic review of the N400 differences. Brain Sci. 2020;10(11):1–45.

21. Koelsch S, Kasper E, Sammler D, Schulze K, Gunter T, Friederici AD. Music, language and meaning: Brain signatures of semantic processing. Nat Neurosci. 2004;7(3):302–7.

22. Miranda RA, Ullman MT. Double dissociation between rules and memory in music : An event-related potential study ☆. 2007;38:331–45.

23. Cui AX, Troje NF, Cuddy LL. Electrophysiological and behavioral indicators of musical knowledge about unfamiliar music. Sci Rep [Internet]. 2022;12(1):1–13. Available from: 10.1038/s41598-021-04211-w

24. Calma-Roddin N, Drury JE. Music, Language, and The N400: ERP Interference Patterns Across Cognitive Domains. Sci Rep [Internet]. 2020;10(1):1–14. Available from: 10.1038/s41598-020-66732-0

25. World Health Organization. Global action plan on the public health response to dementia 2017 - 2025. Geneva: World Health Organization [Internet]. 2017;52. Available from: http://www.who.int/mental_health/neurology/dementia/action_plan_2017_2025/en/

26. Sibbald B, Roberts C. Understanding controlled trials Crossover trials. Bmj. 1998;316(7146):1719–20.

27. Pavlicevic M, Tsiris G, Wood S, Powell H, Graham J, Sanderson R, et al. The ‘ripple effect’: Towards researching improvisational music therapy in dementia care homes. Dementia. 2015;14(5):659–79.

28. Särkämö T, Sihvonen AJ. Golden oldies and silver brains: Deficits, preservation, learning, and rehabilitation effects of music in ageing-related neurological disorders. Cortex. 2018;109(iii):104–23.

29. Folstein MF, Folstein SE, McHugh PR. “Mini-mental state”: A practical method for grading the cognitive state of patients for the clinician. J Psych Res. 1975;12((3)):189–198.

30. Vanstone AD, Wolf M, Poon T, Cuddy LL. Measuring engagement with music: Development of an informant-report questionnaire. Aging Ment Health. 2016;20(5):474– 84.

31. Vanstone AD, Cui AX, Cuddy LL. Using fsQCA to illuminate person attributes of music engagement in Alzheimer’s disease. Music Sci (Lond). 2023;6(November).

32. Coppalle R, Mauger C, Hommet M, Segobin S, Letortu O, De la Sayette V, et al. Recognition-based memory through familiarity assessment in severe Alzheimer’s disease. Brain Cogn [Internet]. 2019;137(December):103639. Available from: 10.1016/j.bandc.2019.10.008

33. Schafer K, Aisen P, Böhm P, Connor D, Desanti S, Gessert D, et al. Administration and Scoring Manual Alzheimer’s Disease Assessment Scale – Cognitive (ADAS-cog) (03/12 modification). Alzheimer’s Disease Cooperative Study. 2012;13:1–34.

34. Liu KY, Schneider LS HR. The need to show minimum clinically important differences in Alzheimer’s disease trials. Lancet Psychiatry. 2021;8:1013–1016.

35. Bell AJ, Sejnowski TJ. The “independent components” of natural scenes are edge filters. Vision Res. 1997 Dec 1;37(23):3327–38.

36. Baker FA, Bloska J, Braat S, Bukowska A, Clark I, Hsu MH, et al. HOMESIDE: Home-based family caregiver-delivered music and reading interventions for people living with dementia: Protocol of a randomised controlled trial. BMJ Open. 2019;9(11):1–11.

37. Fusar-Poli L, Bieleninik Ł, Brondino N, Chen XJ, Gold C. The effect of music therapy on cognitive functions in patients with dementia: a systematic review and meta-analysis. Aging Ment Health [Internet]. 2018;22(9):1097–106. Available from: 10.1080/13607863.2017.1348474

38. Särkämö T, Tervaniemi M, Laitinen S, Numminen A, Kurki M, Johnson JK, et al. Cognitive, emotional, and social benefits of regular musical activities in early dementia: Randomized controlled study. Gerontologist. 2014;54(4):634–50.

39. Balsis S, Benge JF, Lowe DA, Geraci L, Doody RS. How Do Scores on the ADAS-Cog, MMSE, and CDR-SOB Correspond? Clinical Neuropsychologist. 2015;29(7):1002–9.

40. Metcalfe C. The analysis of cross-over trials with baseline measurements. Stat Med. 2010;29(30):3211–8.

41. Norcross JC, Wampold BE. What works for whom: Tailoring psychotherapy to the person. J Clin Psychol. 2011;67(2):127–32.

